# Assessment of genes involved in lysosomal diseases using the ClinGen Clinical Validity framework

**DOI:** 10.1101/2024.08.09.24311755

**Authors:** Emily Groopman, Shruthi Mohan, Amber Waddell, Matheus Wilke, Raquel Fernandez, Meredith Weaver, Hongjie Chen, Hongbin Liu, Deeksha Bali, Heather Baudet, Lorne Clarke, Christina Hung, Rong Mao, Filippo Pinto e Vairo, Lemuel Racacho, Tatiana Yuzyuk, William J Craigen, Jennifer Goldstein

**Author notes:** Corresponding author: Jennifer Goldstein, email address, Mailing address: Department of Genetics UNC-Chapel Hill CB#7264, Chapel Hill, NC 27599-7264.

## Abstract

Lysosomal diseases (LDs) are a heterogeneous group of rare genetic disorders that result in impaired lysosomal function, leading to progressive multiorgan system dysfunction. Accurate diagnosis is paramount to initiating targeted therapies early in the disease process in addition to providing prognostic information and appropriate support for families. In recent years, genomic sequencing technologies have become the first-line approach in the diagnosis of LDs. Understanding the clinical validity of the role of a gene in a disease is critical for the development of genomic technologies, such as which genes to include on next generation sequencing panels, and the interpretation of results from exome and genome sequencing. To this aim, the ClinGen Lysosomal Diseases Gene Curation Expert Panel utilized a semi-quantitative framework incorporating genetic and experimental evidence to assess the clinical validity of the 56 LD-associated genes on the Lysosomal Disease Network’s list. Here, we describe the results, and the key themes and challenges encountered.

## 1. INTRODUCTION

Lysosomal diseases (LDs) are a diverse group of genetic disorders that lead to impaired function of lysosomes. Lysosomes are cellular organelles that calibrate cellular metabolism in response to environmental cues via multiple pathways, including degrading macromolecules delivered via endocytosis, phagocytosis, and autophagy, nutrient sensing, and cellular signaling^1, 2^. Although individually rare, LDs cumulatively occur in approximately 1 in 5,000 to 7,500 births and can result in progressive dysfunction of multiple organ systems, reflecting the importance of lysosomes for metabolic homeostasis^3^. Importantly, for multiple LDs, targeted treatments, including enzyme replacement, substrate reduction therapy, pharmaceutical chaperones, and/or hematopoietic stem cell transplantation, exist, and early initiation of therapy can substantially improve health outcomes^1^. Accordingly, LDs are being included in newborn screening programs in many nations^4^.

Traditionally, diagnosis of LDs relied on clinical symptomatology and assessment of enzymatic activity^5^. However, as rare disorders that frequently have nonspecific and overlapping manifestations (e.g., skeletal anomalies, progressive neurological dysfunction) and span a broad spectrum of clinical severity^3^, LDs can be difficult for physicians to recognize, leading to delayed diagnosis and poor outcomes^5^. Moreover, while deficient activity of the associated enzyme is consistent with an LD, certain variants in LD-associated genes can result in pseudodeficiency, where individuals have deficient enzymatic activity detected by *in vitro* assays but show no clinical signs of disease^5^. In addition, certain LDs result from variants in non-enzymatic lysosomal proteins, and thus are unable to be diagnosed via enzymatic activity assays^5^.

Molecular genetic testing is emerging as a key technology for identification of individuals suspected to have LDs. This approach has the potential to enable early diagnosis and initiation of treatment early in the natural history of disease^3,5^. Genetic testing is increasingly done using next generation sequencing (NGS)-based approaches that assess many genes (e.g., multigene panels) or look across the entire genome (e.g., exome or genome sequencing), and these approaches are often used for individuals who present with nonspecific symptoms that are not distinct to a particular LD^4,6,7^. In the context of such broad-scope genetic testing, knowledge of the clinical validity of the relationship between a given gene and LD is critical^8^.

The Clinical Genome Resource (ClinGen) is a National Institutes of Health (NIH)-funded initiative that aims to define the clinical relevance of genes and variants to empower precision medicine^9^. To advance this goal, ClinGen has developed a semi-quantitative framework to assign clinical validity to gene–disease relationships^10^, which has been successfully implemented by ClinGen Gene Curation Expert Panels (GCEPs) for a variety of disorders^9^. This framework involves the curation of primary published literature to score genetic and experimental evidence to support the assignment of a clinical validity classification (definitive, strong, moderate, limited, disputed, refuted, or no known disease relationship). The ClinGen Lysosomal Diseases Gene Curation Expert Panel (LD GCEP) has applied this framework to systematically assess the clinical validity of the 56 LD-associated genes from the Lysosomal Disease Network (LDN) list (https://lysosomaldiseasenetwork.org/official-list-of-lysosomal-diseases/). Here, we evaluate the genetic and experimental evidence to classify the clinical validity of each of the 56 LDN genes asserted to cause diseases that affect the lysosomes. We discuss the key themes and associated challenges encountered in this endeavor.

## 2. MATERIALS AND METHODS

### 2.1. The ClinGen Lysosomal Diseases Gene Curation Expert Panel

The ClinGen LD GCEP (https://clinicalgenome.org/affiliation/40110/) was assembled by the ClinGen Inborn Errors of Metabolism Clinical Domain Working Group in 2021 to assess the clinical validity of gene-disease relationships for genes involved in lysosomal diseases. This group currently includes 18 individuals from different institutions within the USA and Canada. Seven members of the GCEP are biocurators with training on the ClinGen gene-disease clinical validity framework, and assigned to identify, collect, and assess relevant data from published literature and public databases (such as gnomAD); several of these individuals also have clinical, research, and/or laboratory experience in LDs. Nine members of the GCEP are experts with significant clinical, research, and/or laboratory expertise in LDs who are assigned to review the data collected by the biocurators, to discuss and questions around those findings, and to come to a consensus on the clinical validity classification of a gene-disease pair. Two individuals, one with significant experience in metabolic disorders, coordinate the activities of the group.

### 2.2. List of genes for curation

The main workflow is shown in Fig 1. We began by reviewing the LDN list, which consisted of 56 genes (https://lysosomaldiseasenetwork.org/official-list-of-lysosomal-diseases/). Because LDs are typically multi-systemic conditions, we were aware that some of the gene-disease pairs on the list may already have been curated by other ClinGen GCEPs that focus on phenotypes that are included in LDs such as neurodevelopmental delay (Intellectual Disability-Autism GCEP). Therefore, we surveyed the ClinGen website to determine which of these genes had already been curated. The curated data and clinical validity classifications of genes already curated by other ClinGen GCEPs were reviewed by the LD GCEP to ensure that we agreed with the assessment of the data and final clinical validity classification. For genes that did not already have a curated disease entity published on the ClinGen website, we used the ClinGen Gene Tracker to determine if any other GCEPs planned to curate any of the genes and then contacted those GCEPs to discuss priorities and possible collaboration.

**Figure 1.**
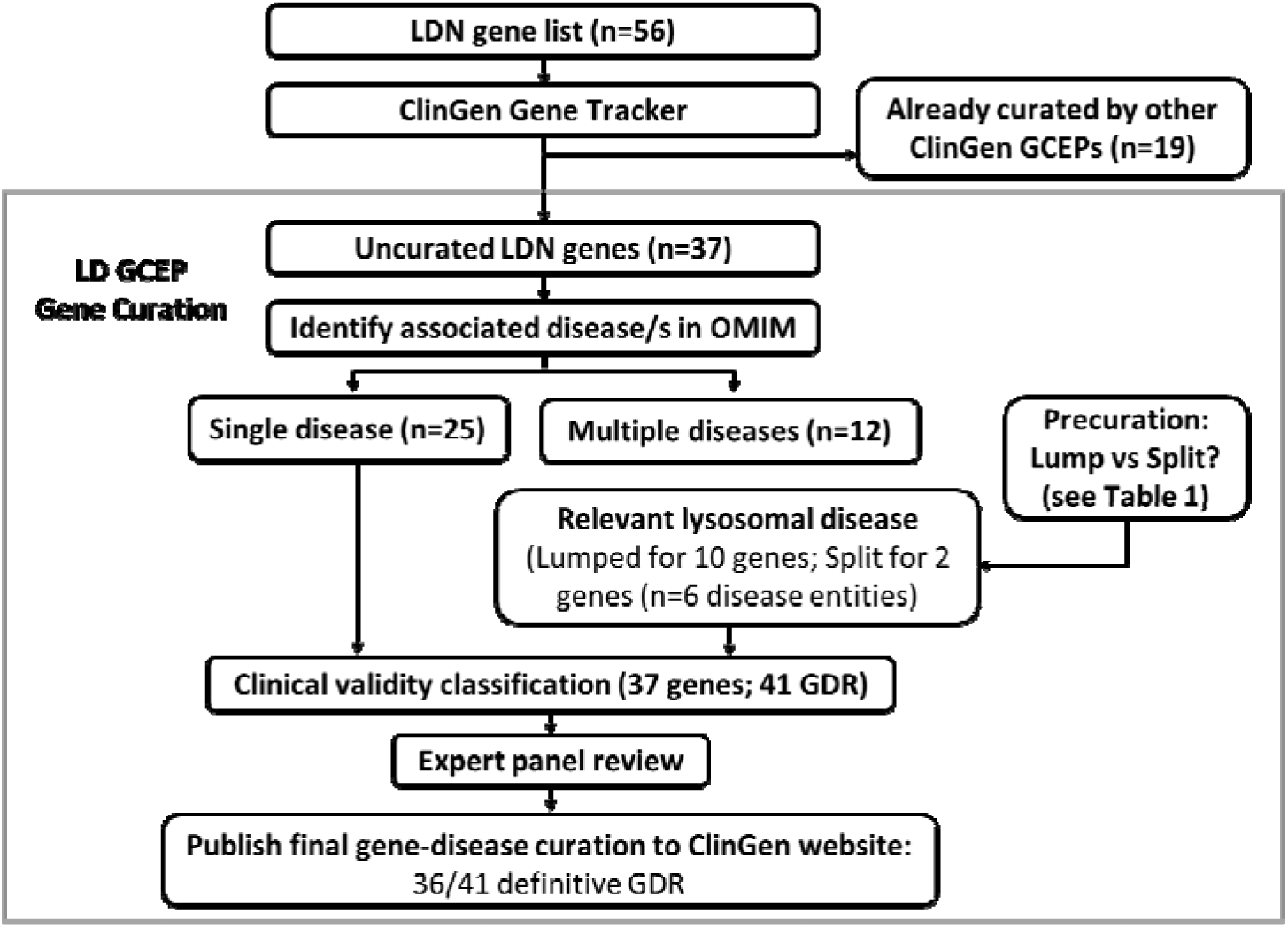
LD GCEP gene curation workflow. Beginning with the LDN “Official List of Lysosomal Diseases’’ (n=56), we found 37 gene-disease pairs that had not been curated and applied the ClinGen Gene-Disease Validity Framework to assess their clinical validity, a process which included: pre-curation and Lumping/Splitting of any genes associated with multiple diseases (n=12; Table 1); full curation of the gene-disease entity; and expert panel review of the curation. Final gene-disease curations are published to the ClinGen website (https://search.clinicalgenome.org/kb/gene-validity).

**Table 1.**
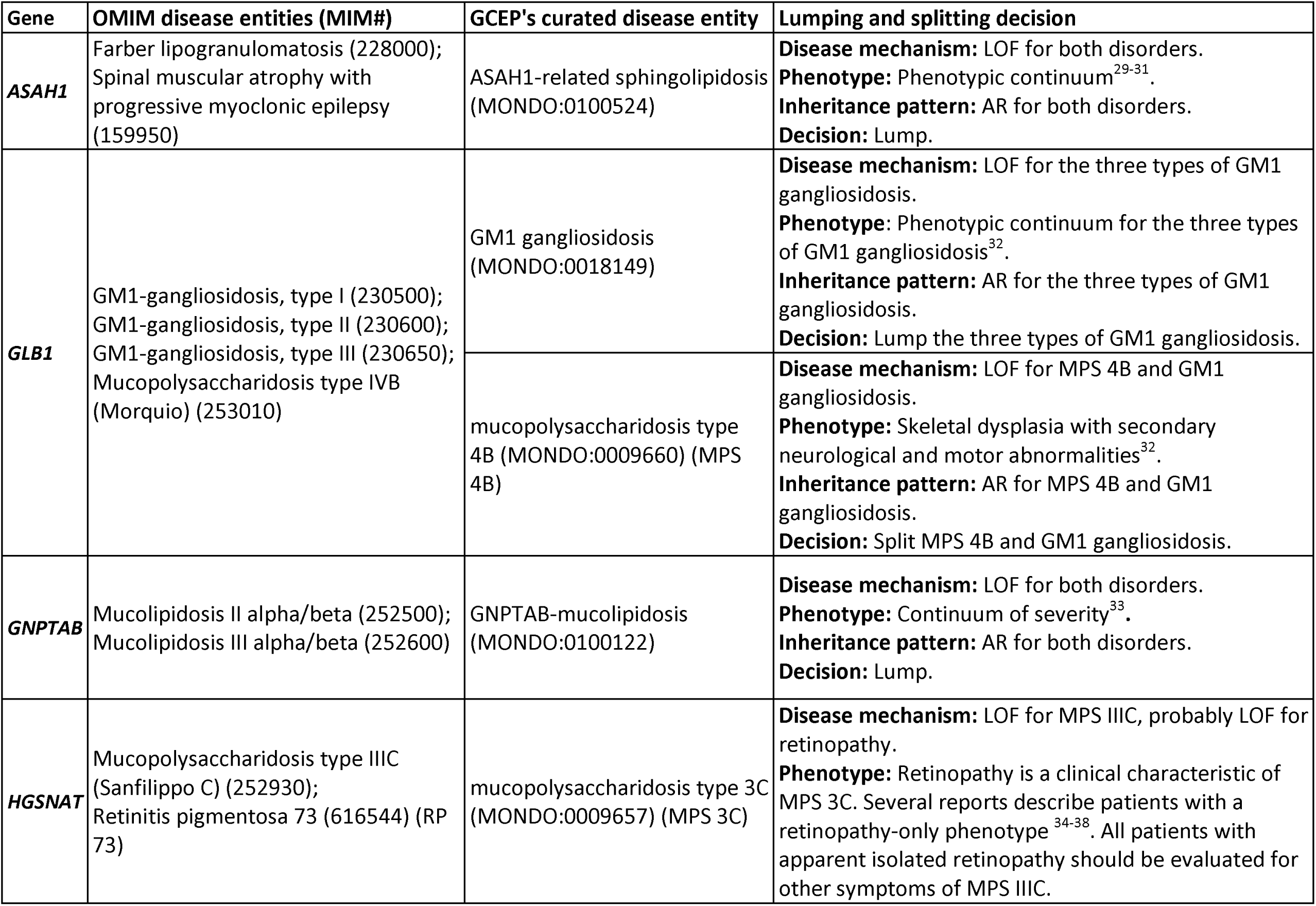
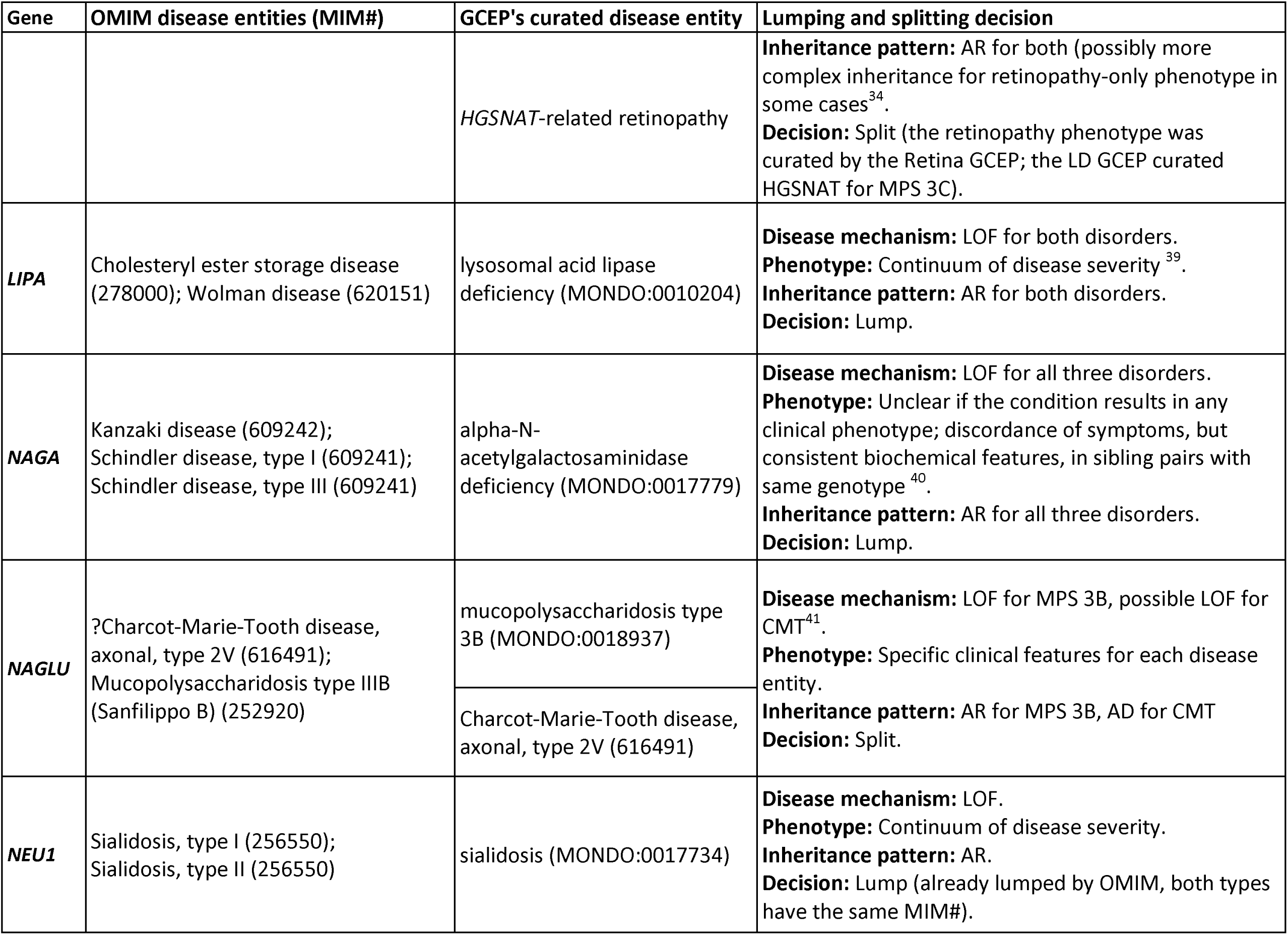
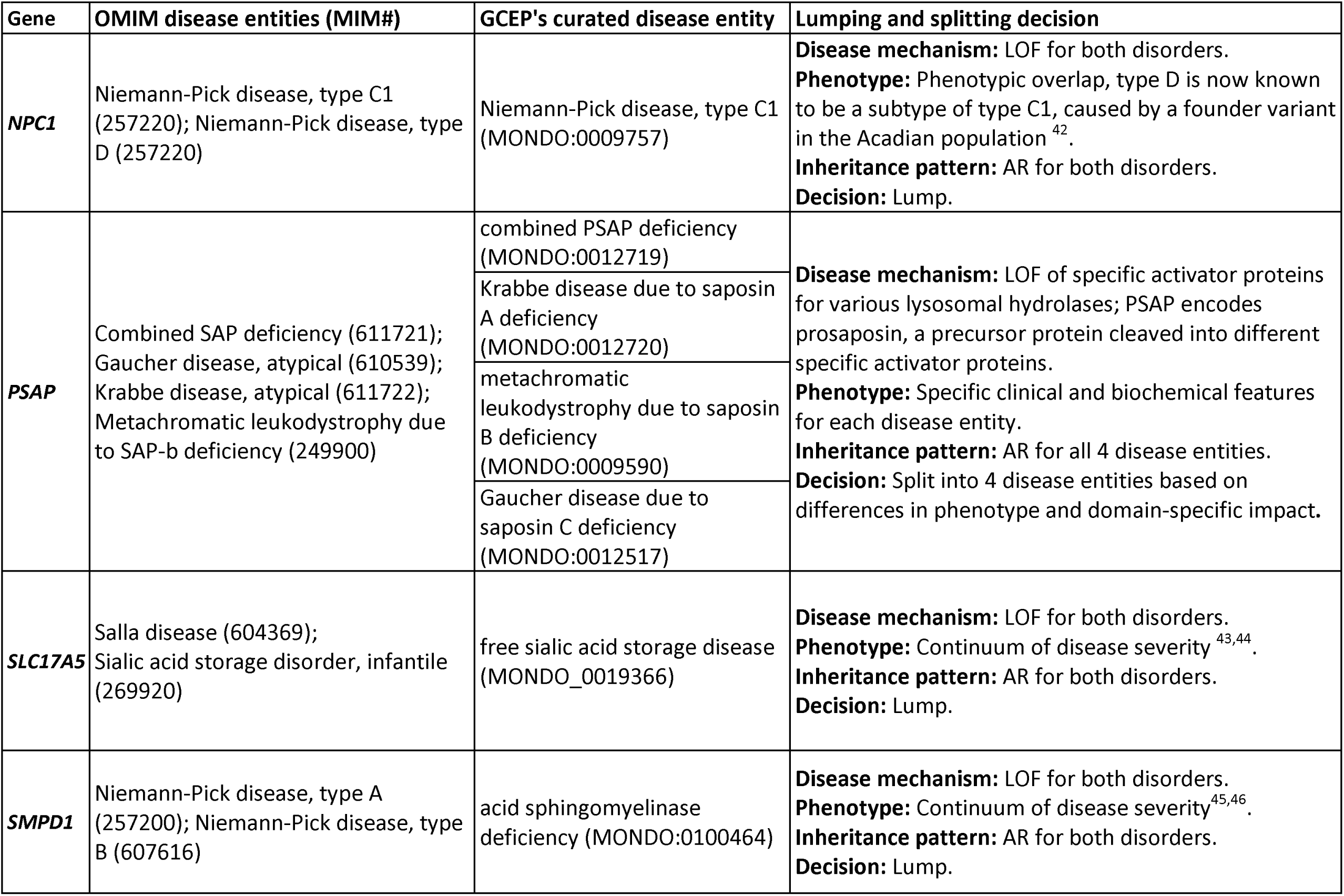
Lumping and splitting, and disease-naming decisions by the LD GCEP for genes with multiple disease entities in OMIM.

| Gene | OMIM disease entities (MIM#) | GCEP's curated disease entity | Lumping and splitting decision |
| --- | --- | --- | --- |
| **ASAH1** | Farber lipogranulomatosis (228000);<br>Spinal muscular atrophy with<br>progressive myoclonic epilepsy<br>(159950) | ASAH1-related sphingolipidosis<br>(MONDO:0100524) | **Disease mechanism:** LOF for both disorders.<br>**Phenotype:** Phenotypic continuum<sup>29-31</sup>.<br>**Inheritance pattern:** AR for both disorders.<br>**Decision:** Lump. |
| **GLB1** | GM1-gangliosidosis, type I (230500);<br>GM1-gangliosidosis, type II (230600);<br>GM1-gangliosidosis, type III (230650);<br>Mucopolysaccharidosis type IVB<br>(Morquio) (253010) | GM1 gangliosidosis<br>(MONDO:0018149) | **Disease mechanism:** LOF for the three types of GM1<br>gangliosidosis.<br>**Phenotype:** Phenotypic continuum for the three types<br>of GM1 gangliosidosis<sup>32</sup>.<br>**Inheritance pattern:** AR for the three types of GM1<br>gangliosidosis.<br>**Decision:** Lump the three types of GM1 gangliosidosis. |
|  |  | mucopolysaccharidosis type<br>4B (MONDO:0009660) (MPS<br>4B) | **Disease mechanism:** LOF for MPS 4B and GM1<br>gangliosidosis.<br>**Phenotype:** Skeletal dysplasia with secondary<br>neurological and motor abnormalities<sup>32</sup>.<br>**Inheritance pattern:** AR for MPS 4B and GM1<br>gangliosidosis.<br>**Decision:** Split MPS 4B and GM1 gangliosidosis. |
| **GNPTAB** | Mucopolipidosis II alpha/beta (252500);<br>Mucopolipidosis III alpha/beta (252600) | GNPTAB-mucopolipidosis<br>(MONDO:0100122) | **Disease mechanism:** LOF for both disorders.<br>**Phenotype:** Continuum of severity<sup>33</sup>.<br>**Inheritance pattern:** AR for both disorders.<br>**Decision:** Lump. |
| **HGSNAT** | Mucopolysaccharidosis type IIIC<br>(Sanfilippo C) (252930);<br>Retinitis pigmentosa 73 (616544) (RP<br>73) | mucopolysaccharidosis type 3C<br>(MONDO:0009657) (MPS 3C) | **Disease mechanism:** LOF for MPS IIIC, probably LOF for<br>retinopathy.<br>**Phenotype:** Retinopathy is a clinical characteristic of<br>MPS 3C. Several reports describe patients with a<br>retinopathy-only phenotype<sup>34-38</sup>. All patients with<br>apparent isolated retinopathy should be evaluated for<br>other symptoms of MPS IIIC. |
|  |  | HGSNAT-related retinopathy | **Inheritance pattern:** AR for both (possibly more complex inheritance for retinopathy-only phenotype in some cases<sup>34</sup>.<br>**Decision:** Split (the retinopathy phenotype was curated by the Retina GCEP; the LD GCEP curated HGSNAT for MPS 3C). |
| **LIPA** | Cholesteryl ester storage disease (278000); Wolman disease (620151) | lysosomal acid lipase deficiency (MONDO:0010204) | **Disease mechanism:** LOF for both disorders.<br>**Phenotype:** Continuum of disease severity<sup>39</sup>.<br>**Inheritance pattern:** AR for both disorders.<br>**Decision:** Lump. |
| **NAGA** | Kanzaki disease (609242); Schindler disease, type I (609241); Schindler disease, type III (609241) | alpha-N-acetylgalactosaminidase deficiency (MONDO:0017779) | **Disease mechanism:** LOF for all three disorders.<br>**Phenotype:** Unclear if the condition results in any clinical phenotype; discordance of symptoms, but consistent biochemical features, in sibling pairs with same genotype<sup>40</sup>.<br>**Inheritance pattern:** AR for all three disorders.<br>**Decision:** Lump. |
| **NAGLU** | ?Charcot-Marie-Tooth disease, axonal, type 2V (616491); Mucopolysaccharidosis type IIIB (Sanfilippo B) (252920) | mucopolysaccharidosis type 3B (MONDO:0018937) | **Disease mechanism:** LOF for MPS 3B, possible LOF for CMT<sup>41</sup>.<br>**Phenotype:** Specific clinical features for each disease entity.<br>**Inheritance pattern:** AR for MPS 3B, AD for CMT<br>**Decision:** Split. |
|  |  | Charcot-Marie-Tooth disease, axonal, type 2V (616491) |  |
| **NEU1** | Sialidosis, type I (256550); Sialidosis, type II (256550) | sialidosis (MONDO:0017734) | **Disease mechanism:** LOF.<br>**Phenotype:** Continuum of disease severity.<br>**Inheritance pattern:** AR.<br>**Decision:** Lump (already lumped by OMIM, both types have the same MIM#). |
| **NPC1** | Niemann-Pick disease, type C1 (257220); Niemann-Pick disease, type D (257220) | Niemann-Pick disease, type C1 (MONDO:0009757) | **Disease mechanism:** LOF for both disorders.<br>**Phenotype:** Phenotypic overlap, type D is now known to be a subtype of type C1, caused by a founder variant in the Acadian population <sup>42</sup>.<br>**Inheritance pattern:** AR for both disorders.<br>**Decision:** Lump. |
| **PSAP** | Combined SAP deficiency (611721); Gaucher disease, atypical (610539); Krabbe disease, atypical (611722); Metachromatic leukodystrophy due to SAP-b deficiency (249900) | combined PSAP deficiency (MONDO:0012719)<br>Krabbe disease due to saposin A deficiency (MONDO:0012720)<br>metachromatic leukodystrophy due to saposin B deficiency (MONDO:0009590)<br>Gaucher disease due to saposin C deficiency (MONDO:0012517) | **Disease mechanism:** LOF of specific activator proteins for various lysosomal hydrolases; PSAP encodes prosaposin, a precursor protein cleaved into different specific activator proteins.<br>**Phenotype:** Specific clinical and biochemical features for each disease entity.<br>**Inheritance pattern:** AR for all 4 disease entities.<br>**Decision:** Split into 4 disease entities based on differences in phenotype and domain-specific impact. |
| **SLC17A5** | Salla disease (604369); Sialic acid storage disorder, infantile (269920) | free sialic acid storage disease (MONDO_0019366) | **Disease mechanism:** LOF for both disorders.<br>**Phenotype:** Continuum of disease severity <sup>43,44</sup>.<br>**Inheritance pattern:** AR for both disorders.<br>**Decision:** Lump. |
| **SMPD1** | Niemann-Pick disease, type A (257200); Niemann-Pick disease, type B (607616) | acid sphingomyelinase deficiency (MONDO:0100464) | **Disease mechanism:** LOF for both disorders.<br>**Phenotype:** Continuum of disease severity <sup>45,46</sup>.<br>**Inheritance pattern:** AR for both disorders.<br>**Decision:** Lump. |

### 2.3. Curation process

Clinical validity curations for all genes on the LDN list were performed according to the ClinGen Gene Disease Clinical Validity framework^10^. All data was entered into ClinGen’s online Gene Curation Interface (GCI). In brief, the ClinGen Clinical Validity framework is a semi-quantitative system that awards points for specific pieces of case-level (genetic) and experimental evidence that support a gene-disease relationship. Once curation of the evidence is complete, the scored evidence is summed to yield a total point score and corresponding clinical validity classification. Definitive and strong clinical validity classifications are achieved by 12 points or more. In addition, definitive classifications must have case-level evidence replicated over a time of three years or more. Moderate and limited classifications are met for 8-12 and 0.1-6 points respectively. No known disease relationship is used as the clinical validity descriptor if there has never been an assertion that variants in the gene are associated with human disease.

Previously asserted gene-disease relationships can also be disputed or refuted based on the degree of conflicting evidence. In addition, as gene-disease validity classifications may change over time (e.g., due to new evidence being published since the original curation), we recommend that readers check the ClinGen website (www.clinicalgenome.org) for the most up-to-date information. Further details and the current Standard Operating Procedure (SOP) are available on the ClinGen website (https://clinicalgenome.org/curation-activities/gene-disease-validity/) and provided in Supplemental file 1)

In the LD GCEP, each curation was performed by a single biocurator with expertise in the ClinGen clinical validity framework. To begin a curation within the GCI, biocurators must first select a gene from the assigned list (using HGNC nomenclature), disease entity for that gene (denoted by a Mondo ID), and inheritance pattern for curation. The ClinGen Lumping and Splitting guidelines^11^ were used to determine the appropriate disease entity, or entities, for curation. For genes assessed by the LD GCEP, all data used to decide whether a disease for a specific gene should be “lumped” together for curation, or “split” into different curations were discussed during the GCEP’s regular online meetings. After collecting and scoring relevant evidence, biocurators assigned a provisional classification for each gene-disease relationship that was then reviewed by experts in the group. Evidence for gene-disease relationships with a provisional clinical validity classification less than definitive or curations for which a biocurator had questions were reviewed on a group conference call during which the biocurator would present all curated data and scoring to the group. While a quorum of 2 was required, typically, at least 5 experts, with experience ranging from clinical, research, and molecular diagnostics, were present on each call and would come to a consensus on the final clinical validity classification. Definitive provisional classifications were reviewed offline by at least two experts. Once approved by two or more experts, the clinical validity classification and associated data for each curation were published to the ClinGen website (www.clinicalgenome.org), where they are openly accessible.

## 3. RESULTS AND DISCUSSION

### 3.1. Genes for curation

Based upon review of the ClinGen website, 17 genes on the LDN list had already been curated by other ClinGen GCEPs. This was not surprising considering that many LDs include phenotypes under the purview of other GCEPs, such as neurodevelopmental delay (Intellectual Disability-Autism (ID-Autism) GCEP). Members of LD GCEP reviewed the curated data and clinical validity classifications for these disease-disease pairs and agreed with the final classifications. Of note, 5 of these genes had previously been curated for the General GCEP by an individual with significant experience in lysosomal diseases who is now a biocurator for the LD GCEP. Of the remaining 39 genes, 30 were on other GCEPs lists for curation; 28 of those genes were not considered to be high priority for curation by those other GCEPs and were, therefore, transferred to the LD GCEP’s list. The remaining two genes (*CTNS, CLCN5*) were high priority for the Tubulopathy and Glomerulopathy GCEPs, given that renal disease is the primary feature associated with these genes. These genes were therefore curated by the Tubulopathy and Glomerulopathy GCEPs (Figure 1). One of the LD GCEP biocurators is also a member of these GCEPs, thus ensuring consistency in the evaluation of the data.

### 3.2. Determining the disease entity for curation

Of the 37 LDN genes on our list, 12 genes were associated with more than one condition in OMIM, and one gene (*SLC38A9*) had no disease entity listed in OMIM. We followed the ClinGen lumping and splitting guidelines to determine the appropriate disease entity, or entities, for those genes^11^ (Table 1). We chose to lump together any disease entities that represented different severities and ages of onset for similar phenotypic characteristics and therefore represented a clinical spectrum rather than distinct disease entities. For example, the disease entities Niemann-Pick disease, type A, and Niemann-Pick disease, type B, were lumped as acid sphingomyelinase deficiency (ASMD) because these two disorders represent a clinical spectrum with the same mechanism of disease (loss of function), and the inheritance pattern for both conditions is the same (autosomal recessive). When necessary, we requested a new Mondo disease identifier via the Mondo GitHub (https://github.com/monarch-initiative/mondo/issues) for lumped disease entities. For example, acid sphingomyelinase deficiency was not previously listed in Mondo. This term has since been added based on our request (MONDO:0100464), with the child terms, already present in Mondo, of Niemann-Pick disease type A (MONDO:0009756), and Niemann-Pick disease type B (MONDO:0011871). While there is utility in using more specific disease descriptors when discussing phenotypic features, prognosis, and treatment with patients and families, ClinGen recommends lumping disease entities across a phenotypic spectrum, for the purposes of gene-disease clinical validity assessment, when there is no clear indication to split diseases into separate entities. Lumping across a clinical spectrum is common for metabolic disorders, most of which are recessive with phenotype at least partly dependent on the impact of a combination of two alleles, and where genotype/phenotype correlation is not always known.

For four genes curated by the LD GCEP (*GLB1*, *HGSNAT*, *NAGLU*, *PSAP*), there were substantial differences in the phenotypes of their respective disease entities, and thus they were split for curation(Table 1). For example, after detailed discussions with members of the Retina GCEP, we split the disease entities for *HGSNAT* into mucopolysaccharidosis type IIIC (MPS IIIC; MIM# 252930) and retinitis pigmentosa 73 (MIM# 616544). The LD GCEP proceeded to curate *HGSNAT* for MPS IIIC, and the Retina GCEP has curated *HGSNAT* for retinitis pigmentosa 73 (Table 1). We plan to revisit the *HGSNAT* data again every 2 years to ensure that no additional data has emerged that may lead us to lump these conditions together, such as reports of additional features developing in MPS IIIC in patients originally reported to have isolated retinopathy,

### 3.3. Clinical validity classifications: LDN list

Of the 37 LDN genes on our list, 33 were given a clinical validity classification of definitive for at least one disease entity (Figure 2, Supplemental Table 1). For the 19 LDN genes curated by other GCEPs, all were definitive for at least one disease entity (Supplemental Table 2). Therefore, in total, 52/56 (92.9%) genes on the LDN list were definitive for one or more disease entities.

**Figure 2.**
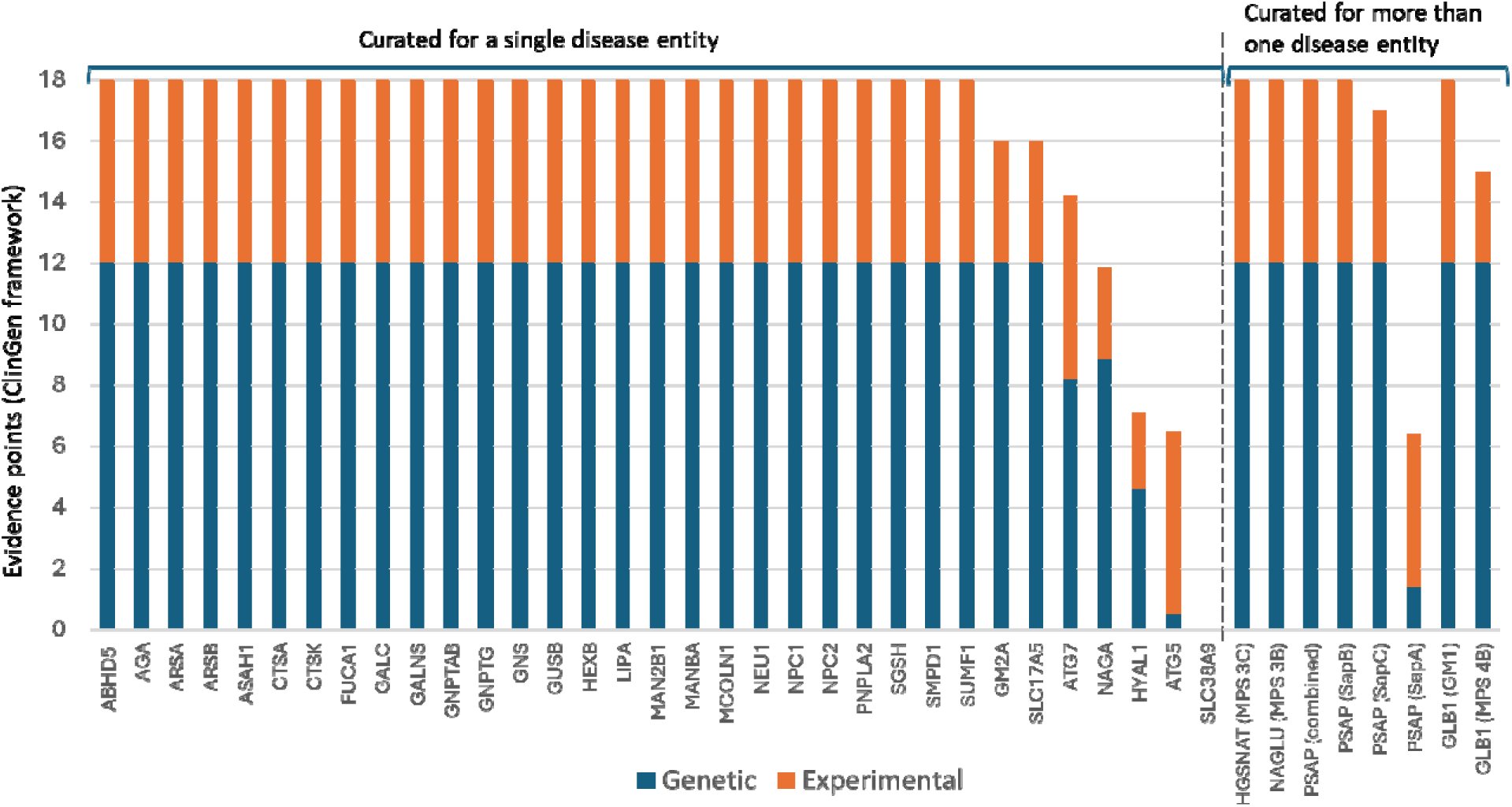
ClinGen Gene-Disease Clinical Validity evidence points for genes on the LDN list curated by the LD GCEP. 37 genes on the LDN list, that had not already been curated by other ClinGen GCEPs, underwent gene-disease clinical validity classification. 33 of those genes were curated for a single disease entity, while 4 were curated for split disease entities (Table 1, Supplemental Table 1). The additional 19 genes on the LDN list have been classified by other ClinGen GCEPs; all are definitive for at least one disease entity (Supplemental Table 2).

Including the genes with split disease entities, the LD GCEP curated 41 gene disease-relationships for the LDN list (Table 1, Supplemental Table 1). This includes 4 diseases for *PSAP*, and 2 for *GLB1*. In total, 36 of the 41 gene-disease relationships curated were definitive (Supplemental Table 1). For all but one of the definitive gene-disease relationships, the genetic evidence met or exceeded 12 points, which is the maximum points that can be counted for genetic evidence. The one definitive gene-disease relationship which did not have maximum genetic evidence was *NAGA*-alpha-N-acetylgalactosaminidase deficiency (MONDO:0017779). At the time of curation, nine probands had been reported in the literature; eight with biallelic missense variants and one homozygous for a nonsense variant. The total genetic evidence score was 8.85 points. The addition of experimental evidence (biochemical function and non-human animal model) brought the classification to definitive (11.85 points, rounded to 12; with approval of “definitive” by the GCEP members). Of interest, variants in *NAGA* have been associated with Schindler disease, type I (MIM# 609241; NAGA deficiency type I), Schindler disease, type III (MIM# 609241; NAGA deficiency type III), and Kanzaki disease (MIM# 609242; also known as Schindler disease, type II; NAGA deficiency type II). However, clinically unaffected family members have been reported with the same genotype and enzyme deficiency^12,13^. The variability in clinical phenotype from asymptomatic to neurological manifestations does not diminish the definitive relationship between *NAGA* and “alpha-N-acetylgalactosaminidase deficiency“; all reported individuals with biallelic variants in the gene had the enzyme deficiency; other factors, such as genetic modifiers, have been suggested to influence the genotype-phenotype correlation^13^. However, while the relationship between variants in *NAGA* and the biochemical phenotype of alpha-N-acetylgalactosaminidase deficiency is definitive, the clinical impact of alpha-N-acetylgalactosaminidase deficiency is unclear at the current time.

All the definitive gene-disease relationships on the LDN list that were curated by the LD GCEP (n=36) had supportive experimental evidence. This included evidence indicating that the biochemical function of the gene product is consistent with phenotypic features in patients with the disease, and all had non-human animal models that recapitulate features of the disease. Additional evidence curated for some genes included protein interaction, for those genes encoding subunits of a multi-subunit complex or enzyme; rescue in humans for genes causing conditions for which enzyme replacement therapy is available; and rescue experiments in animal models or cultured cells. In total 34 (94%) of the definitive gene disease relationship met or exceeded the 6-point maximum for experimental evidence and the remaining 2 (6%) were given 4 points.

Four of the 37 LDN genes on our list were found to not be definitively associated with their respective LD: *ATG7* (strong), *HYAL1* (moderate), *ATG5* (limited), and *SLC38A9* (no known disease relationship) (Figure 2; Supplemental Table 1). *ATG7* had a strong classification for the associated disease entity, spinocerebellar ataxia, autosomal recessive 31, a neurodevelopmental disorder with multisystem involvement and impaired autophagy^14^. While *ATG7* was described in the LDN list as a “non-disease protein,” biallelic *ATG7* variants were reported in 5 families with features of complex neurodevelopmental disorder with brain, muscle, and endocrine involvement^14^, yielding 8.2 points for genetic evidence. This genetic evidence, along with experimental evidence including the biochemical function of the protein^15^ and a mouse model recapitulating the human disease phenotype^16^, led to a classification of strong (total 14.2 points). Although the total score exceeded the 12-point cutoff for a definitive gene-disease association, the ClinGen curation guidelines^10^ require replication of the genetic evidence (i.e., further reported cases) over 3 or more years in order for the classification to be definitive. This requirement was not met for *ATG7*. *HYAL1* was curated for mucopolysaccharidosis type IX (MONDO:0011093). At the time of curation, there were only 2 published probands, one of whom had two affected siblings^17–19^, yielding 4.6 points of genetic evidence. Experimental evidence (2.5 points) included the expression pattern of the gene^18^ and an animal model^20^. A total of 7.1 points was thus given, yielding a moderate classification. *ATG5* was curated for spinocerebellar ataxia, autosomal recessive 25. Only one family, with two affected siblings, had been published at the time of the curation, leading to 0.5 points being given for genetic evidence^21^. The gene-disease relationship was supported by the biochemical function of the gene product^22^ and a drosophila model in which the phenotype was rescued by the wild type gene^21^ (6 points experimental evidence). Based on the level of supportive evidence, the gene disease relationship was classified as limited. While the points awarded summed to 6.5, borderline between limited (0.1-6 points) and moderate (7-11 points), the GCEP members agreed on a limited classification for this gene-disease relationship based on a limited number of reported cases. *SLC38A9* was given a classification of “no known disease relationship” because, based on our literature review, there were no reports describing individuals with a clinical phenotype caused by variants in the gene. Based on published experimental data, there is evidence suggesting a role for *SLC38A9* in lysosomal disease. *SLC38A9* encodes “solute carrier family 38, member 9”, a lysosomal amino acid transporter protein involved in regulating the mechanistic target of rapamycin complex 1 (mTORC1)^23–25^. SLC38A9 physically interacts with NPC intracellular cholesterol transporter 1^25^, a protein encoded by the NPC1 gene, which is definitively associated with Niemann-Pick disease type C1. HEK293T cells harboring *SLC38A9* variants lacking key amino acid residues have impaired amino acid-mediated signaling of mTORC1^24^; and abnormal amino acid metabolism was observed in a *SLC38A9* knockout zebrafish model^26^. While there is currently “no known disease relationship” for SLC38A9, ClinGen adds the additional tag of “animal model only” to reflect this evidence.

### 3.4. ClinVar classifications based on clinical validity

The importance of the clinical validity classification is reflected in the classifications of variants submitted to ClinVar (data accessed March 2, 2024) (Supplemental Table 3). This is consistent with current recommendations^28^ that genes with a clinical validity classification of limited or below, in general, should not be included on diagnostic panels or reported from exome or genome analyses. *ATG7*, has relatively few variants reported in ClinVar (48 total variants, 6 pathogenic), which may reflect that the first cases were in 2021 (spinocerebellar ataxia, autosomal recessive 31). While *HYAL1* was found to have moderate clinical validity, cases were first reported in 1999^18^; accordingly, there are 349 total variants, including 34 pathogenic and 2 likely pathogenic, in ClinVar. *ATG5* and *SLC38A9*, which were respectively curated as limited and no human disease relationship, have no pathogenic or likely pathogenic variants submitted to ClinVar by clinical diagnostic laboratories, indicating the uncertainty of the clinical significance of variants in these genes based on limited or no clinical validity evidence (Supplementary Table 3). Based on ClinGen’s recommendations for recuration (https://clinicalgenome.org/site/assets/files/2164/clingen_standard_gene-disease_validity_recuration_procedures_v1.pdf), the gene-disease relationships that do not have a definitive classification will be re-evaluated in the future in order to look for and evaluate any new evidence that could impact the classification.

### 3.5. Challenges and key takeaways

Molecular genetic testing has the capacity to expedite diagnosis of LDs, supporting early therapeutic interventions and improved outcomes for individuals and their families^3,5^. With NGS-based multigene panels or genome-wide sequencing increasingly used as first-line techniques, genetic testing for LD involves analyzing many genes, which may vary in the strength to which they are associated with their respective LD. To accurately interpret the pathogenicity of the genetic variants identified, one must first assess the clinical validity of the relationship between the given gene with its disease entity^8^.

Here, we applied a systematic framework integrating genetic and experimental evidence^10^ to classify the clinical validity of the 56 proposed LD genes from the LDN. 52 of these 56 (92.9%) genes were definitively associated with at least one LD, supporting their inclusion in the genetic evaluation of suspected LD. Of the remaining 4 genes, two had limited (*ATG5*) or no evidence (*SLC38A9*) for association with LD; in ClinVar, these genes had a high burden of VUS, highlighting the analytic challenges posed by genes with uncertain clinical validity and the need for cautious interpretation if including them in diagnostic genetic evaluation.

Our experience highlights several major themes and associated challenges for classifying the clinical validity of proposed LD genes. Twelve of the 37 LDN genes curated by the LD GCEP were associated with multiple disease entities, and thereby required precuration to determine whether these disease entities should be lumped together within a single disease spectrum, or split as separate conditions (Table 1). As LDs frequently have clinically heterogeneous and variably severe presentations^3^, it can be difficult to distinguish whether a given set of disease entities fall within a single disease spectrum or instead are distinct disorders. To address this challenge, we applied the ClinGen Lumping/Splitting heuristic to assess the overlap in disease mechanism and clinical manifestations; for eight genes, there was sufficient overlap to lump the disease entities, whereas for the remaining four genes, the molecular genetics and/or clinical features of the disease entities differed enough to merit them being split. As illustrated by the example of acid sphingomyelinase deficiency, our work on lumping and splitting altered the categorization of the associated disorders in MONDO, emphasizing how systematic curation can provide new insight regarding gene-disease relationships. With the advent of LD patient registries^27^, we anticipate that the understanding of the phenotypic spectrum of LDs will continue to grow, thereby informing lumping/splitting for LDs.

Classifying the clinical validity of a proposed LD gene requires evaluating both the genetic and experimental evidence supporting their association. As the majority of LDN genes had been first reported in association with their LD several decades ago, there was robust genetic and experimental evidence available, conferring a definitive classification. However, for several of the more recently reported genes, there were fewer published cases available, contributing to non-definitive classifications with their LDs. Given the rarity of LDs, it can be difficult to interpret whether a relative dearth of published cases argues against the gene-disease association or instead reflects the fact that individuals with the given genetic disease are exceedingly rare. In addition to continued reexamination of the published literature to identify newly reported cases, data-sharing initiatives such as Matchmaker Exchange^28^ can help address this issue via connecting individuals with similar phenotypes who have variants in a given candidate gene. Moreover, for our initial curation efforts, we focused on genes that were on the LDN list. This approach may have biased our curations towards definitive and strong gene-disease relationships.

## 4. CONCLUSION

In our study, we chose to focus on the 56 LDN reported genes, reflecting its role as a consensus LD gene list. Our work provides a well-annotated and openly accessible resource of classified LD genes; however, as an initial curation effort, our approach does not capture all candidate LD genes. Future work will include the development and validation of a set of criteria to identify novel candidate LD genes in order to prioritize our curation efforts. In addition, our classifications are limited by the available genetic and experimental evidence at the time of classification; thus, we plan to revisit the LD genes classified and reassess their clinical validity based on evaluation of any newly available evidence. Nonetheless, our work offers a systematic framework that can be applied to classify the clinical validity of LD genes, thereby increasing our understanding of the genetic and phenotypic spectrum of LD and supporting early diagnosis and therapeutic interventions for individuals with LD.

## Data Availability

All data produced in the present study are available upon reasonable request to the authors

## Abbreviations

GCEP: Gene Curation Expert Panel
LD: Lysosomal Disease
LDN: Lysosomal Disease Network

## Acknowledgements

We are grateful to the ClinGen Inborn Errors of Metabolism Clinical Domain Working Group for their insightful comments and support through this work, the ClinGen Gene Curation Working Group for their review of the manuscript, and the Stanford ClinGen Informatics team for support.

## CRediT authorship contribution statement

E.G. – conceptualization, formal analysis, investigation, writing (original draft), writing (review and editing); S.M. - investigation, writing (review and editing); A.W. - investigation, writing (review and editing); M.V.M.B.W. - investigation, writing (review and editing); R.F. - project administration; M.W. - conceptualization, project administration; H.C. - investigation; H.L. - investigation, D.B. - verification, writing (review and editing); H.B.- verification, writing (review and editing); L.C. - verification, writing (review and editing); C.H. – verification, writing (review and editing), R.M. - conceptualization, verification, writing (review and editing); F.P.e.V- conceptualization, verification, writing (review and editing); L.R.- verification; T.Y. - verification, writing (review and editing); W.J.C.- conceptualization, verification, writing (review and editing); J.L.G. – conceptualization, formal analysis, investigation, supervision, writing (original draft), writing (review and editing).

